# Near infrared spectroscopy during non-ischaemic handgrip exercise reveals divergent muscle oxygenation signatures in mitochondrial disease and sarcopenia

**DOI:** 10.64898/2026.10.21.26364901

**Authors:** Helen Devine, Anderson Brito Da Silva, Susan J. Hillman, Callum G. Brownstein, Sinead Smith, Kevin Rao, Sarah Pickett, Doug Turnbull, Roger G Whittaker, Christopher Hurst, Miles D Witham, Avan A Sayer, Thomas C. Bulea, Rachel Cooper, Karen Suetterlin

**Author notes:** Corresponding author: Karen Suetterlin.

## Abstract

**Background:** Mitochondrial dysfunction is a hallmark of ageing and is implicated in the pathogenesis of sarcopenia. Genetically confirmed mitochondrial disease provides a biologically definitive model of impaired oxidative phosphorylation against which novel functional assessments can be validated. We investigated whether a brief, submaximal, non-ischaemic intermittent isometric handgrip exercise protocol combined with Near Infra-Red Spectroscopy (NIRS) could detect distinct muscle oxygenation signatures in mitochondrial disease and sarcopenia.

**Methods:** Adults with genetically confirmed mitochondrial disease (n=10, 7 female) and group-matched controls (n=19, 11 female), underwent assessment of maximal grip force, handgrip relaxation kinetics and a 3-minute intermittent submaximal isometric handgrip test (40% maximal voluntary contraction, 1Hz contraction/relaxation) with continuous force recording at 2000Hz and simultaneous NIRS. Findings were compared with 23 community-dwelling older adults from the Muscle Ageing and Sarcopenia Study (MASS) Lifecourse cohort (n=23, 13 female) including seven with probable or confirmed sarcopenia according to European Working Group on Sarcopenia in Older People (EWGSOP2) criteria.

**Results:** Participants with mitochondrial disease demonstrated an *attenuated* reduction in tissue saturation index (TSI) during exercise compared with controls (TSI %·s150-180s: −59.9±94.5 vs −175.1±128.4; p=0.02). Total haemoglobin (THb) responses did not differ between groups (p=0.10). In contrast, participants with probable or confirmed sarcopenia showed *exaggerated* reduction in TSI compared with non-sarcopenic participants (TSI %·s150-180s: −256.5±146.0 vs −121.2±95.2; p=0.05), while THb responses remained similar. The association between sarcopenia and TSI differed by sex (interaction coefficient −240, p=0.016), with greater TSI reductions observed in males with sarcopenia. Differences in TSI between mitochondrial disease and control groups were independent of age and sex. No statistically significant associations were observed between THb responses and age, sex or force production.

**Conclusions:** A brief, non-ischaemic handgrip exercise protocol combined with NIRS identified divergent muscle oxygenation responses in mitochondrial disease and sarcopenia. The *attenuated* TSI response observed in genetically confirmed mitochondrial disease is consistent with impaired oxygen utilisation secondary to defective oxidative phosphorylation, whereas the *exaggerated* TSI reduction observed in sarcopenia suggests a distinct physiological mechanism, potentially involving impaired oxygen delivery, reduced muscle efficiency, or both. This portable and non-invasive approach has significant potential as a physiological biomarker for mechanistic studies and clinical trials in ageing and neuromuscular disease. More broadly, our findings demonstrate how biologically defined rare diseases can provide a powerful framework for interpreting mechanisms in common heterogeneous age-related conditions.

## Introduction

Sarcopenia, defined as the accelerated loss of skeletal muscle mass and function, is a major contributor to disability, hospitalisation and premature mortality in older adults^1,2^. Muscle weakness is a core component in many of the most widely utilised diagnostic criteria for sarcopenia, including the European Working Group on Sarcopenia in Older People (EWGSOP2) consensus, with confirmation based on reduced muscle quantity or quality and severity determined by impaired physical performance^3^. Grip strength, assessed using handheld dynamometry, has consequently become a widely used screening and diagnostic tool because it is rapid, inexpensive, and feasible across healthcare settings^4^. However, whilst grip strength provides valuable prognostic information, it captures only peak force during a brief isometric contraction and offers limited insight into the biological mechanisms underlying muscle dysfunction. This is important because sarcopenia is likely to be biologically heterogeneous and approaches capable of identifying distinct physiological perturbations may improve mechanistic understanding and support stratification in future clinical trials.

Mitochondrial dysfunction is a hallmark of ageing^5^ and is implicated in the pathogenesis of sarcopenia^6^. Exercise increases skeletal muscle ATP turnover up to 100-fold and under oxidative conditions, mitochondria provide the vast majority of this ATP. Repeated submaximal contraction-relaxation protocols may therefore reveal abnormalities in skeletal muscle bioenergetics that are not apparent from a single maximal voluntary contraction. However, clinically accessible approaches for assessing muscle mitochondrial function remain limited. The current gold-standard for non-invasive assessment of oxidative capacity is the phosphocreatine recovery curve measured using ^31^P-magnetic resonance spectroscopy (^31^P-MRS). However, its wider application is limited by cost, availability, and technical requirements and Magnetic Resonance Imaging (MRI) is contraindicated in some patient groups. Near-infrared spectroscopy (NIRS) provides a portable, non-invasive alternative approach for assessing skeletal muscle oxygenation. Muscle oxygenation reflects the balance between oxygen delivery and oxygen utilisation, and impaired muscle oxidative metabolism may therefore alter the extent of muscle oxygen extraction during exercise. Recovery kinetics of muscle oxygen extraction measured using NIRS (NIRS mV̇O_2_) correlate closely with phosphocreatine recovery curves measured using ^31^P-magnetic resonance spectroscopy (MRS)^7^ supporting its use as an indirect assessment of skeletal muscle oxidative function. During exercise, impaired oxygen utilisation associated with mitochondrial dysfunction would therefore be expected to produce less muscle deoxygenation and an *attenuated* reduction in tissue saturation index (TSI).

Genetically confirmed mitochondrial dysfunction provides a biologically defined human model of impaired oxidative phosphorylation in which the ability of NIRS to detect meaningful physiological abnormalities can be tested. Previous studies have demonstrated impaired muscle oxygen extraction in mitochondrial myopathy using NIRS during cycle ergometry testing^8^. However, this requires bulky specialist equipment to perform and is less suitable for translation into clinical or community settings, where a handgrip test would be preferable. Whilst a handgrip NIRS mV̇O2 study did not distinguish individuals with mitochondrial myopathies from controls^9^, a handgrip exercise-to-exhaustion protocol combined with NIRS demonstrated high sensitivity for detecting impaired muscle oxygen extraction, with patients with genetically confirmed mitochondrial disease showing an attenuated decline in deoxyhaemaglobin signal (HHb) during exercise compared with healthy controls^10,11^. This protocol required arterial occlusion with a pneumatic cuff to achieve steady-state HHb for calibration, which may limit tolerability and wider applicability. Impaired oxygen extraction has previously been demonstrated in mitochondrial myopathy patients using an intermittent, submaximal, isometric handgrip exercise task combined with repeated venous blood oxygenation measurements^12^. If the same physiological abnormality could be detected non-invasively using NIRS, this would provide a simpler, more cost-effective, and more clinically applicable means of measuring muscle oxygenation without the need for arterial occlusion or specialist exercise equipment.

Establishing that a protocol can detect abnormalities in genetically confirmed mitochondrial disease is an important first step before applying the same approach to more biologically heterogeneous conditions such as sarcopenia. NIRS has been less extensively used to characterise sarcopenia. A recent study demonstrated altered tissue oxygenation during fatiguing lower-limb exercise in older adults with sarcopenia but required laboratory-based exercise equipment. It remains unclear whether a brief, clinically accessible exercise paradigm can detect altered muscle oxygenation in sarcopenia and whether the physiological response resembles that seen in primary mitochondrial dysfunction.

In this study, we combined continuous high frequency digital dynamometry with a brief, non-ischaemic intermittent and submaximal isometric handgrip exercise protocol with simultaneous NIRS assessment. We first tested whether the protocol could detect altered muscle oxygenation in individuals with genetically confirmed mitochondrial disease to provide a biologically defined reference phenotype of impaired oxidative phosphorylation. We then applied an identical protocol in participants recruited to the Muscle Ageing and Sarcopenia Study (MASS) Lifecourse^13,14^ who were classified according to EWGSOP2 criteria^3^. This allowed us to assess whether EWGSOP2-defined probable or confirmed sarcopenia was associated with a similar physiological phenotype of impaired oxidative phosphorylation, or a distinct physiological pattern.

## Methods

### Participants

This work integrates data from two observational studies. The first cohort comprised adults with genetically confirmed mitochondrial disease and exercise intolerance (score≥1) recruited from the MitoCohort^15^ and controls without exercise intolerance or neuromuscular diagnosis recruited via a Newcastle University healthy volunteer mailing list. Participants with mitochondrial disease had additional phenotyping data available including genotype, Newcastle Mitochondrial Disease Adult Scale (NMDAS) scores^16^ and, where available, heteroplasmy levels (**Table 1**). The mitochondrial dysfunction study participants were recruited under the ‘Developing Non- or Minimally Invasive Measures of Mitochondrial Dysfunction’ study, approved by the Southwest Central Bristol Research Ethics Committee (23/SW/0052), while the MitoCohort (The Mitochondrial Disease Patient Cohort: A Natural History Study and Patient Registry) was approved by the Newcastle and North Tyneside Research Ethics Committee (14/SS/0020).

**Table 1.** Characteristics of participants with genetically confirmed mitochondrial disease ordered by TSI %·s 150-180s. NMDAS = Newcastle Mitochondrial Disease Adult Scale. GLTEQ = Godin Leisure-Time Exercise Questionnaire. TSI %·s 150-180s: Integrated TSI (%·s) values calculated for the last 30-second interval of the 3-minute hand-grip exercise test and normalised to a mean TSI during the stable pre-exercise period. *analysis of Cytochrome C Oxidase deficient fibres found > 85% heteroplasmy in 2 separate skeletal muscle biopsies.

| Patient | Age category (years) | Sex | Genetics | GLTE Q score | NMDAS scaled score | Corrected heteroplasmy | Tissue | TSI %·s 150-180s | Myopathy | Ptosis | CPEO | Diabetes | Stroke-like episodes |
| --- | --- | --- | --- | --- | --- | --- | --- | --- | --- | --- | --- | --- | --- |
| 1 | 56-60 | F | Single deletion | 24 | 28 | - | - | 39.41 | Yes | Yes | Yes | No | No |
| 2 | 51-55 | M | m.12271T>C | 21 | 24.7 | 0.1* | Muscle | 23.48 | Yes | Yes | Yes | Yes | No |
| 3 | 31-35 | F | m.3243A>G | 25 | 14.5 | 0.9 | Urine | 0.88 | No | No | No | No | No |
| 4 | 56-60 | F | m.3243A>G | 0 | 17.2 | 0.44 | Blood | -9.28 | No | No | No | No | No |
| 5 | 81-85 | F | TWINKLE (PEO1) | 24 | 23.2 | - | - | -39.04 | No | Yes | Yes | No | No |
| 6 | 51-55 | M | m.3243A>G | 3 | 53.5 | 0.84 | Urine | -51.39 | Yes | No | No | Yes | No |
| 7 | 66-70 | F | m.3243A>G | 11 | 12.8 | 0.7 | Blood | -76.6 | No | No | No | Yes | No |
| 8 | 36-40 | F | NDUFB3 | 0 | 18.6 | - | - | -197.33 | Yes | No | No | No | No |
| 9 | 51-55 | M | m.3243A>G | 56 | 30.1 | 0.66 | Muscle | -229.36 | Yes | No | No | Yes | Yes |

The second cohort comprises a subset of 23 participants from MASS Lifecourse who completed the ‘New Measures’ sub study and as part of their baseline assessment underwent the tests described below^14^. MASS Lifecourse exclusion criteria were pregnancy, diabetes mellitus, immunosuppressant, anticoagulant or antiplatelet medications (except for aspirin for primary prevention of cardiovascular disease which could be suspended), receiving palliative care, housebound, living in a residential or nursing home. Participants were classified according to EWGSOP2 sarcopenia criteria. This cohort enabled assessment of whether physiological signatures identified in mitochondrial disease were also associated with sarcopenia. The MASS Lifecourse study was approved by the North East Newcastle & North Tyneside 1 Research Ethics Committee (18/NE/0220).

All participants provided written informed consent prior to study participation.

### Revised HandClench Relaxometer (rHCR)

The revised HandClench Relaxometer (rHCR) is adapted from an original design described previously^17^ to create a fully portable hand grip force measurement system operating on a microcontroller (Raspberry Pi 4; OS 11). The system incorporates a speaker and touchscreen display providing automated visual and auditory cues synchronized with continuous force recording at 2000 Hz. Embedded software enables task-specific force recording, real-time visual feedback of grip force during data collection and subsequent analysis of contraction and relaxation kinetics.

To complete grip force tasks, participants were seated with the elbow of the dominant hand positioned at approximately 90°, and the forearm secured to ensure consistent joint position throughout testing (**Figure 1A**). The HCR force transducer was placed in the palm of the dominant hand and centred on the middle finger (**Figure 1a**, **1b**). Maximal voluntary contraction (MVC) and relaxation time were measured using the rHCR.

**Figure 1.**
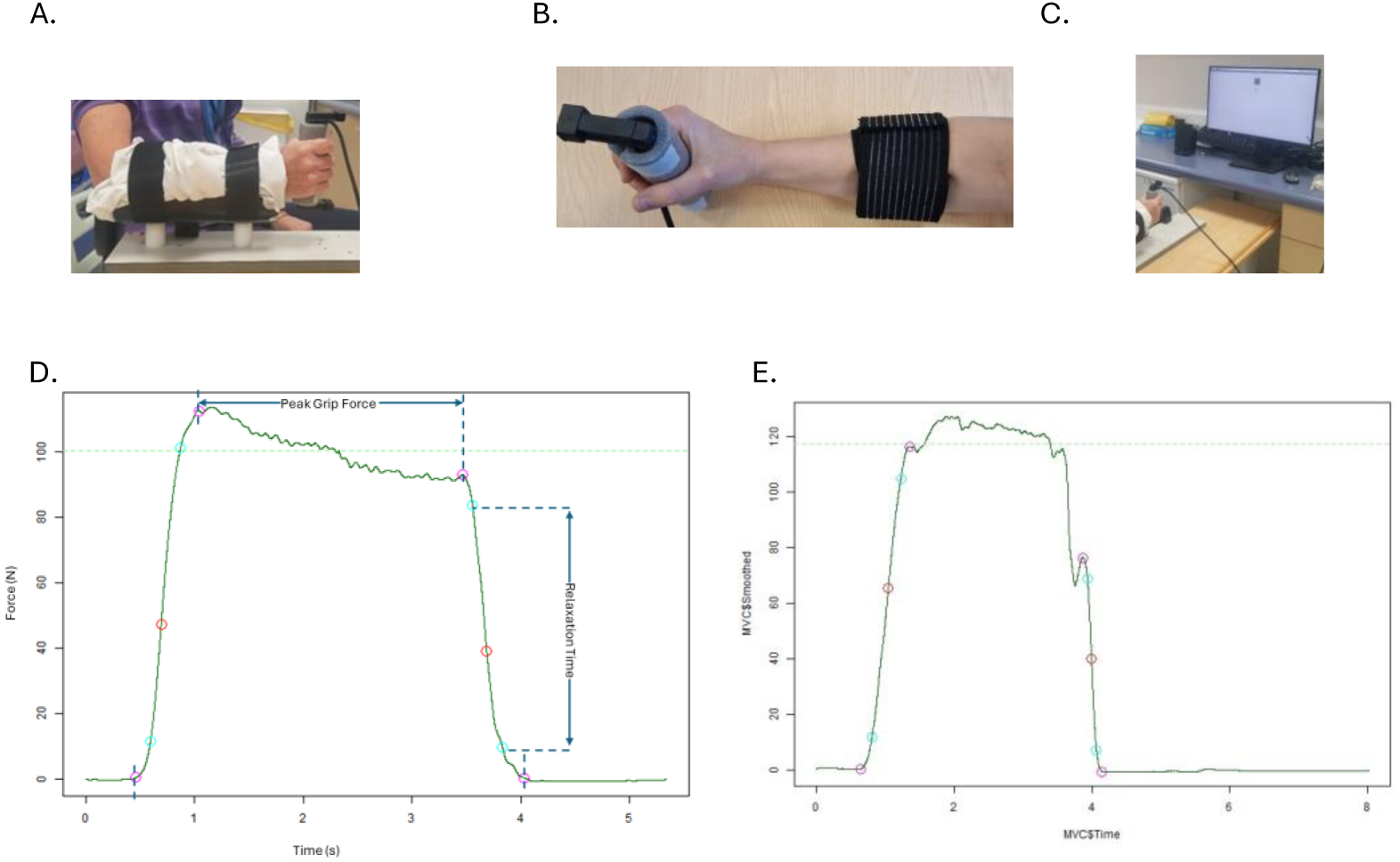
rHCR and NIRs setup. a. The elbow was positioned at approximately 90% and forearm secured in cushioned brace using Velcro with Artinis Portamon device in situ (see b). b. The Artinis Portamon NIRs device was positioned 1/3 of the distance between the medial olecranon and the pisiform bone and medial/lateral positioning optimised to point of maximal muscle bulk on grip squeeze and secured with dark velcroid fabric. c. The participant was positioned in front of a monitor displaying the visual cue – a coloured square. d. An example rHCR trace. Four key transition points (pink circles) were defined by identifying when the gradient decreased to 5% of its peak magnitude before and after the positive and negative extrema. These points corresponded to (i) initiation and (ii) termination of the onset phase, and (iii) initiation and (iv) termination of the relaxation phase. The mean force between termination of the onset phase and initiation of the relaxation phase was used to calculate the peak grip force. The relaxation time was calculated as the time from 90% to 10% force on the descending limb. e..Relaxation phase ‘blip’.

Two force protocols were performed. The first assessed MVC and muscle relaxation kinetics. The second was a repeated hand grip exercise performed during simultaneous NIRS assessment to characterise muscle oxygenation during intermittent submaximal isometric contraction.

### Maximal Voluntary Contraction (MVC)

Participants performed three maximal grip contractions with approximately 30 seconds of rest between attempts. MVC was defined as the highest force recorded across all attempts and expressed in Newtons (N). Where force successively increased across the three trials, one or more additional MVCs were performed until MVC plateaued or declined to ensure the participant had achieved peak grip force.

### 180 second intermittent hand grip exercise

Participants then completed an 85 second familiarisation run at least 15 minutes before an 180 second intermittent handgrip exercise protocol. The handgrip exercise protocol consisted of alternating 1-second isometric contractions at 40% MVC and 1-second relaxation periods^12^.

Automated auditory cues standardised contraction timing whilst visual feedback was used to guide force output within 35-45% of MVC. The display showed a coloured square indicating whether force was within the target range (green for 35-45% MVC), above target range (red for >45% MVC) or below target range (grey for <35% MVC) (**Figure 1c**). Participants were instructed to synchronise hand contraction and relaxation with the auditory cue and try to maintain force within the target range whilst contracting.

### Near infrared spectroscopy (NIRS) and ultrasound

Skeletal muscle oxygenation during exercise was assessed using a portable wireless near-infrared spectroscopy (NIRS) device (PortaMon, Artinis, The Netherlands). The probe was positioned at one-third of the distance between the medial olecranon and the pisiform bone (**Figure 1b**). Probe placement was refined by adjusting medially or laterally to position the device over the point of maximal muscle bulk during hand-clenching to maximise signal quality and reproducibility.

A baseline resting period preceded the exercise protocol, during which participants were instructed to relax and remain as still as possible while holding the rHCR force transducer. Continuous NIRS signals were recorded throughout baseline, exercise, and recovery phases.

Following removal of the NIRS device, subcutaneous tissue thickness at the NIRs measurement site was calculated using a Philips Lumify handheld ultrasound system (Philips Healthcare, Amsterdam, The Netherlands) with an L12-4 linear array transducer (4-12 MHz).

### MVC waveform processing and analysis

Force waveform processing for the MVC task was performed in R using the signal, zoo, and tidyverse packages. Raw force signals were filtered using a bidirectional 40 Hz low-pass Butterworth filter and smoothed with a three-sample moving average. The first derivative was then calculated and smoothed (221-sample moving average) to generate a gradient waveform. Local maxima and minima of this waveform identified the steepest onset and relaxation phases (**Figure 1d**). Four transition points were empirically defined where the gradient fell to 5% of peak magnitude before and after these extrema, marking initiation and termination of contraction and relaxation (pink circles, **Figure 1d**). Peak grip force was calculated as the mean force between the end of contraction and the beginning of relaxation. Relaxation time was defined as the interval from 90% to 10% force on the descending limb (**Figure 1d**).

### MVC quality control

Waveforms with shape irregularities (“blips”) that prevented clear identification of contraction onset, peak force, or relaxation phases (**Figure 1e**) were excluded from analysis. Prespecified blip detection thresholds that were empirically defined were used to identify waveform irregularities during the peak contraction and relaxation phases. During the first 10% of the peak squeeze phase, waveforms were excluded if the minimum force deviated by >2% or the maximum force by >10% of the force at peak squeeze initiation. During the final 10% of the peak squeeze phase, waveforms were excluded if the minimum force was >2% or the maximum force >10% of the force at relaxation initiation. Blips during the relaxation phase were identified if the maximum force in the 0.5 second period following relaxation termination exceeded 5% of the total force decrease during relaxation (**Figure 1d**). The waveform with highest peak grip force that passed all quality criteria was included in subsequent analyses.

### NIRS and force processing during intermittent handgrip exercise

A custom MATLAB application was developed for the integrated analysis of NIRS signals and rHCR output during the 180 second intermittent handgrip exercise protocol. The application is freely available through GitHub (https://github.com/nab221/NIRS_HCR).

Tissue Saturation Index (TSI) was derived using spatially resolved spectroscopy and provides an absolute measure of local tissue oxygenation beneath the sensor. TSI was calculated as the proportion of oxygenated haemoglobin (O₂Hb) relative to total haemoglobin (THb), where THb represents the sum of O_2_Hb and deoxygenated Hb (HHb):

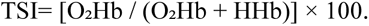

NIRS-derived TSI and THb signals were normalised to a resting baseline, defined as the mean TSI/THb during a stable pre-exercise period of at least 15 seconds. Integrated TSI (%·s) and THb (µM·s) values were calculated for the last 30-second interval of the 180 second hand-grip exercise test (TSI %·s 150-180s and THb µM·s 150-180s respectively). rHCR force output was time-aligned with the NIRS signal allowing total force over the 180 second exercise test and within each 30-second interval to be calculated.

### NIRS quality control

NIRS measures were included if there was ≥15 seconds of stable baseline TSI signal prior to exercise and clear evidence of exercise onset and offset (indicated via ∼1 Hz saw-tooth fluctuation in signal corresponding to intermittent muscle contraction). Data quality assessment was performed independently by a blinded assessor with review and decision by consensus with a second blinded assessor if there was concern over data quality. NIRS measures not meeting quality criteria were excluded from further analysis.

### Physical Activity Assessment

Participants in the “Mitochondrial Dysfunction” study completed the Godin Leisure-Time Exercise Questionnaire (GLTEQ), a validated, self-reported measure of weekly leisure-time physical activity^18^. Participants in MASS Lifecourse completed a Rapid Assessment of Physical Activity (RAPA) questionnaire and wore a GeneActiv accelerometer on their wrist for 7-days prior to the study visit^14^. Time spent participating in moderate-to-vigorous physical activity (MVPA), defined as average daily duration in minutes of activity greater than 100 milligravity units (mg) was calculated using the R package GGIR^19^.

### Additional MASS Lifecourse Measures

In addition to the measures described above, MASS Lifecourse participants underwent a range of additional assessments including grip strength, measured using a Jamar handheld dynamometer^20,21^, five times chair sit to stand (5xSTS), gait speed, height, weight, Dual-energy X-ray Absorptiometry (DXA) and MRI of thigh musculature using a 3T Philips Achieva MRI scanner (Best, NL) as previously described^14^. These measures allowed muscle strength, physical performance, muscle quantity and quality to be characterised and probable, confirmed and severe sarcopenia to be identified according to the EWGSOP2 criteria.

### Statistical Analyses

Data were assessed for normality using the Shapiro–Wilk test. Between-group comparisons were performed using Welch’s t-test or the Mann–Whitney U test, as appropriate. For comparisons involving more than two groups, one- or two-way ANOVA or the Kruskal–Wallis test was used. Linear regression models were used to examine associations between TSI or THb, peak grip force and total force exerted during the 180 second handgrip exercise. Models were stratified by sex a priori to account for the possibility of sex-specific physiological responses and adjusted for age, a known determinant of skeletal muscle function and performance. Statistical analyses were conducted using OriginPro or R, with p < 0.05 considered statistically significant. Data are presented as mean with standard deviation (sd) unless otherwise stated.

## Results

### Participant Characteristics

#### Genetically confirmed mitochondrial disease cohort

A total of 9 participants with genetically confirmed mitochondrial disease (7 female, 2 male) and 19 control participants (11 female, 8 male) were included in analyses; two additional participants were excluded - one participant with mitochondrial disease had a NIRS signal that failed quality control and one control had a diagnosis of chronic fatigue syndrome. The mitochondrial cohort included a range of genetically confirmed mitochondrial diseases (**Table 1**), with five of the nine participants carrying the m.3243A>G variant. Genotype, heteroplasmy levels (where available), and Newcastle Mitochondrial Disease Adult Scale (NMDAS) myopathy and exercise intolerance scores for individual participants are presented in Table 1. There was no statistically significant difference between groups in age (54.9±18.1 vs 55.2±14.8years; p=0.97) or sex distribution (p = 0.52). Participants with mitochondrial disease reported lower habitual physical activity than controls, as assessed by the Godin Leisure-Time Exercise Questionnaire scores (18.6 ± 16.7 vs 48.9 ± 26.2; p<0.01).

#### MASS Lifecourse cohort

Twenty-three participants (13 female, 10 male) from the MASS Lifecourse new measures sub study were included. Mean age was 69.0±9.1years and mean BMI was 25.8±4.0kg/m². Six participants (4 male, 2 female) met EWGSOP2 criteria for probable sarcopenia based on 5xSTS duration exceeding 15 seconds (range 15.16–40.99 seconds). One female participant met EWGSOP2 criteria for confirmed sarcopenia based on 5xSTS >15 seconds, grip strength <16kg and DXA appendicular skeletal muscle mass <15kg. There was no statistically significant difference in age between participants with probable or confirmed sarcopenia and those without sarcopenia (72 ± 8.2 years Sarc Vs 67 ± 9.2 years control, p=0.20).

#### Non-ischaemic handgrip exercise detects altered muscle oxygenation in mitochondrial disease

There was no statistically significant difference in peak grip force between males with mitochondrial disease and male controls (137.8±59.0 Newtons (N) Mito Vs 201.3±69.1N Controls, p=0.18; **Figure 2a**). However, there was a statistically significant difference in peak grip force between females with mitochondrial disease compared with female controls (77.6±16.7N Mito Vs 148.0 ± 59.8N Controls, p= 0.001, Figure 2a). There was no significant difference in relaxation time between males or females with mitochondrial disease compared with their respective control groups (**Figure 2b**).

**Figure 2.**
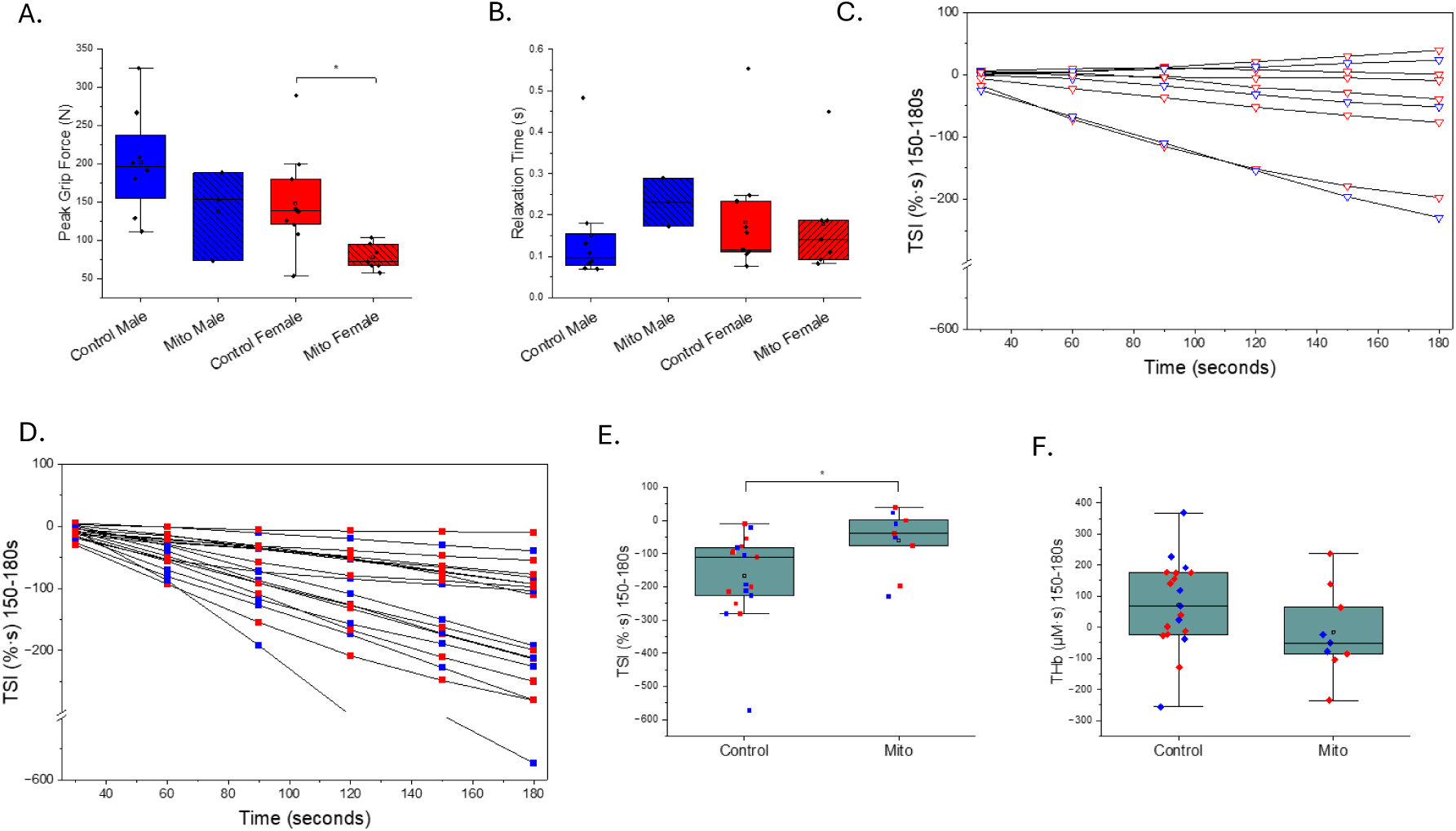
The effect of genetically confirmed mitochondrial dysfunction. a. The rHCR derived peak grip force. b. The rHCR derived Relaxation Time. c. The integrated TSI (%·s) signal over the 180 second hand grip exercise test in participants with genetically confirmed mitochondrial disease, males = blue, females = red. d. The integrated TSI (%·s) signal over the 180 second hand grip exercise test in control participants males = blue, females = red. e. Integrated TSI (%·s) for the last 30 seconds of the 180 second hand grip exercise test. f. Integrated THb (µL·s) for the last 30 seconds of the 180 second hand grip exercise test. Box-and-whisker plots (a.b.d.e) show the mean (empty square). median (centre line), interquartile range (IQR, box), and the most extreme values within 1.5 × IQR (whiskers). Individual data points are shown (blue = male, red =female).

When examining whether the non-ischaemic handgrip protocol identified differences in muscle oxygenation during exercise, two of nine participants with mitochondrial disease demonstrated an increase in TSI during the final 30 seconds of exercise (TSI %·s 150-180s) relative to baseline (**Figure 2c**). This was not observed in any control participant from either the mitochondrial dysfunction study or MASS Lifecourse (**Figure 2d**). Overall, the TSI %·s 150-180s was *attenuated* in participants with mitochondrial disease compared with controls (TSI %·s 150-180s: −59.9±94.5 Vs −175.1±128.4, p=0.02; **Figure 2e**). Differences in TSI %·s (150-180 s) between genotypes were independent of age and sex. A significant association between TSI %·s (150-180 s) and peak grip force or total rHCR force recorded during the exercise protocol was observed when all participants were included (p = 0.02, 0.03 respectively), but this relationship was no longer statistically significant (p = 0.3 for both) following exclusion of a single extreme outlier (male control participant; **Figure 2d,e**)

THb µM·s 150-180s did not differ between groups (78.5±144.2 Vs-15.3±141.4 µM·s, p=0.1, **Figure 2f**) and there were no associations between THb µM·s 150-180s and age, sex, peak grip force or total rHCR force recorded during the exercise protocol. There was no difference in ultrasound-measured forearm subcutaneous tissue thickness between participants with and without mitochondrial disease (0.3±0.2 vs 0.4±0.2cm; p=0.34, two-tailed t-test)

To explore whether the altered TSI response reflected clinical or molecular disease severity, we examined its relationship with NMDAS scores, exercise intolerance, habitual physical activity and corrected heteroplasmy level^22,23^ (Table 1). No clear association was observed between TSI %·s 150-180s and NMDAS total scores, NMDAS exercise intolerance sub scores, or GLTEQ scores. Heteroplasmy data were available for 6 participants (Table 1). No consistent relationship was identified between TSI values and corrected heteroplasmy level^22^ in any tissue.

#### Sarcopenia is associated with an exaggerated reduction in TSI during handgrip exercise

Having identified an attenuated TSI response in genetically confirmed mitochondrial disease we next examined whether sarcopenia was associated with a similar pattern of altered muscle oxygenation. The 6 MASS Lifecourse participants with probable sarcopenia based on prolonged 5xSTS (4 males, 2 females) and the 1 female MASS Lifecourse participant with confirmed sarcopenia were compared with the 16 participants without evidence of sarcopenia.

We first examined handgrip performance and relaxation kinetics. Males with probable sarcopenia had no statistically significant difference in peak grip force when compared with males without sarcopenia (211.7±43.4Newtons Sarc Vs 176.9±38.6 Newtons control, p=0.20, **Figure 3a**) but had *prolonged* hand grip relaxation time (0.15±0.06 seconds Sarc Vs 0.08 ±0.008 seconds control, p=0.02, **Figure 3b**). In contrast, females with probable or confirmed sarcopenia tended to have lower grip strength than females without sarcopenia, (84.8±15.6 Newtons Sarc Vs 108.5±18.2 Newtons control, p=0.09, **Figure 3a**) and had shorter handgrip relaxation time (0.09±0.02 seconds Sarc Vs 0.17±0.07 control p=0.007, **Figure 3b**).

**Figure 3.**
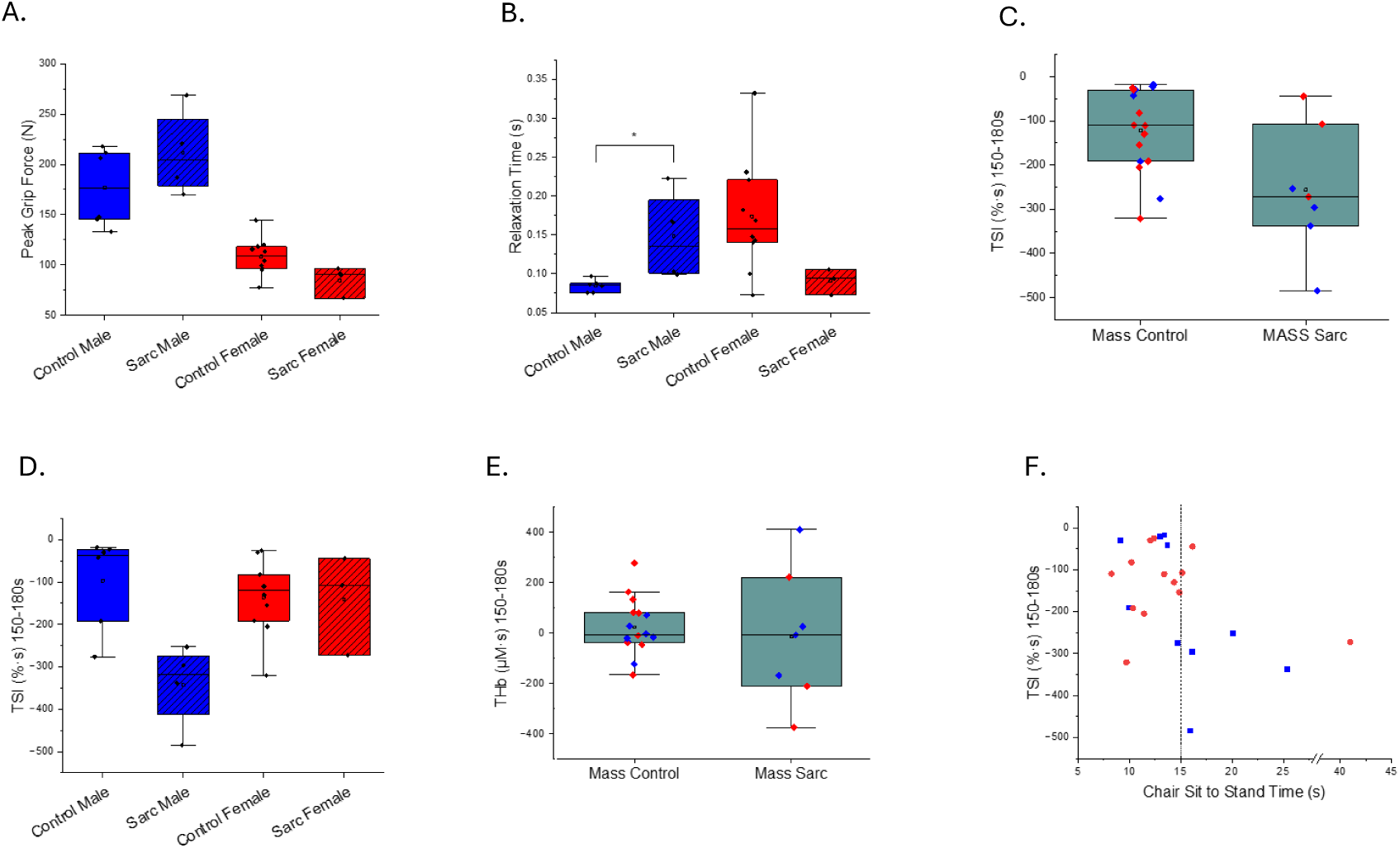
Sarcopenia a. The rHCR derived peak grip force in males and females according to sarcopenia status. b. The rHCR derived Relaxation Time in males and females according to sarcopenia status. c. Integrated TSI (%·s) for the last 30 seconds of the 180 second hand grip exercise test according to sarcopenia. d. Sex-stratified integrated TSI (%·s) for the last 30 seconds of the 180 second hand grip exercise test according to sarcopenia status e. Integrated THb (µL·s) for the last 30 seconds of the 180 second hand grip exercise test according to sarcopenia status. f. Integrated TSI (%·s) for the last 30 seconds of the 180 second hand grip exercise test plotted according to 5xchair sit to stand time for all 23 MASS Lifecourse participants. Box-and-whisker plots (a,b,c,d,e) show the mean (empty square). median (centre line), interquartile range (IQR, box), and the most extreme values within 1.5 × IQR (whiskers). Individual data points are shown (blue = male, red =female).

Next, we explored muscle oxygenation during the same non-ischaemic handgrip protocol. In contrast to participants with genetically confirmed mitochondrial disease, participants with probable or confirmed sarcopenia demonstrated *exaggerated* reduction in TSI compared with those without sarcopenia (TSI %·s 150-180s −256.5±146.0 Vs −121.2±95.2; p=0.05, **Figure 3c**). The effect of sarcopenia differed significantly between sexes (coefficient −240, p = 0.016), indicating a greater reduction in TSI in males with sarcopenia than in females (**Figure 3d**).

THb µM·s 150-180s responses did not differ significantly between participants with and without probable sarcopenia (**Figure 3e**) and there was no evidence of a sex-specific association. There was also no association between rHCR force over the 180 second exercise protocol and either TSI %·s 150-180s or THb µM·s 150-180s.

To explore whether the altered TSI response could be explained by differences in body composition, muscle characteristics or physical activity, we examined these in the MASS Lifecourse participants. There was no difference in BMI between participants with and without probable or confirmed sarcopenia (25.3 ± 4.0 kg/m^2^ Sarc Vs 26.0 ± 4.4 kg/m^2^ Control,p=0.62) nor was there any difference in ultrasound measured forearm subcutaneous tissue thickness (0.5cm±0.6 Sarc Vs 0.3±0.3 Control, p=0.6). Likewise, MRI-derived measures of thigh muscle composition were similar between groups, with no differences observed in muscle volume (5.5 ± 2.1 L units Sarc Vs 5.6 ± 1.6 L Control, p = 0.65), fat fraction (10.9 ± 1.0 % Sarc Vs 10.5 ± 3.0 % Control, p = 0.71), or cross-sectional area (left:134.0 ± 37.8 cm^2^ Sarc Vs 130.2 ± 26.0 cm^2^ Control, p = 0.87; right:133.7 ± 38.8 cm^2^ Sarc Vs 129.3 ± 25.7 cm^2^ Control, p = 0.92).

Participants with probable or confirmed sarcopenia were less physically active with lower accelerometer-derived moderate-to-vigorous-activity (MVPA) than those without sarcopenia (81±35.2 Vs 127±41.8 minutes, p=0.02). Self-reported physical activity was also lower (RAPA1: 6 vs 6.5; p=0.05; RAPA2: 0 vs 2.5; p=0.06). There also appeared to be a positive correlation between chair sit to stand time and TSI %·s 150-180s although this was not statistically significant (**Figure 3f**, p=0.08).

#### Association of age and sex with handgrip and NIRS measures

As age and sex are potential influences on skeletal muscle physiology, we examined their associations with handgrip and NIRS measures in the combined 35 participants (24 female) without genetically confirmed mitochondrial disease or evidence of sarcopenia (19 Mitochondrial Dysfunction study controls and 16 non-sarcopenic MASS Lifecourse participants).

Males had greater peak grip force (195 ±54Newtons Vs 123±47.8Newtons, p<0.001, **Figure 4a**) and faster grip relaxation time (0.13±0.01seconds Vs 0.17±0.1seconds, p=0.02, **Figure 4b**) than females. Increasing age was associated with lower peak grip force in males (coefficient −2.1, p=0.005) but not females (coefficient −1.1, p=0.1, **Figure 5a**). In females, increasing age was associated with a small reduction in relaxation time (coefficient - 0.005, p=0.001, **Figure 5b**). In contrast to the differences observed in grip performance, there was no evidence of a sex difference in TSI %·s 150-180s (−163.6±152%·s males Vs −135.2 ±85.3 females, p=0.5, **Figure 4c**) or THb µM·s 150-180s (42.4±154 µM·s males Vs 53.3 ± 114 µM·s females, p=0.96, **Figure 4d**). Age was not associated with TSI or THb responses in females, while an apparent age-TSI association in males was attributable to a single outlier and was not evident after exclusion (**Figure 4c,d**).

**Figure 4.**
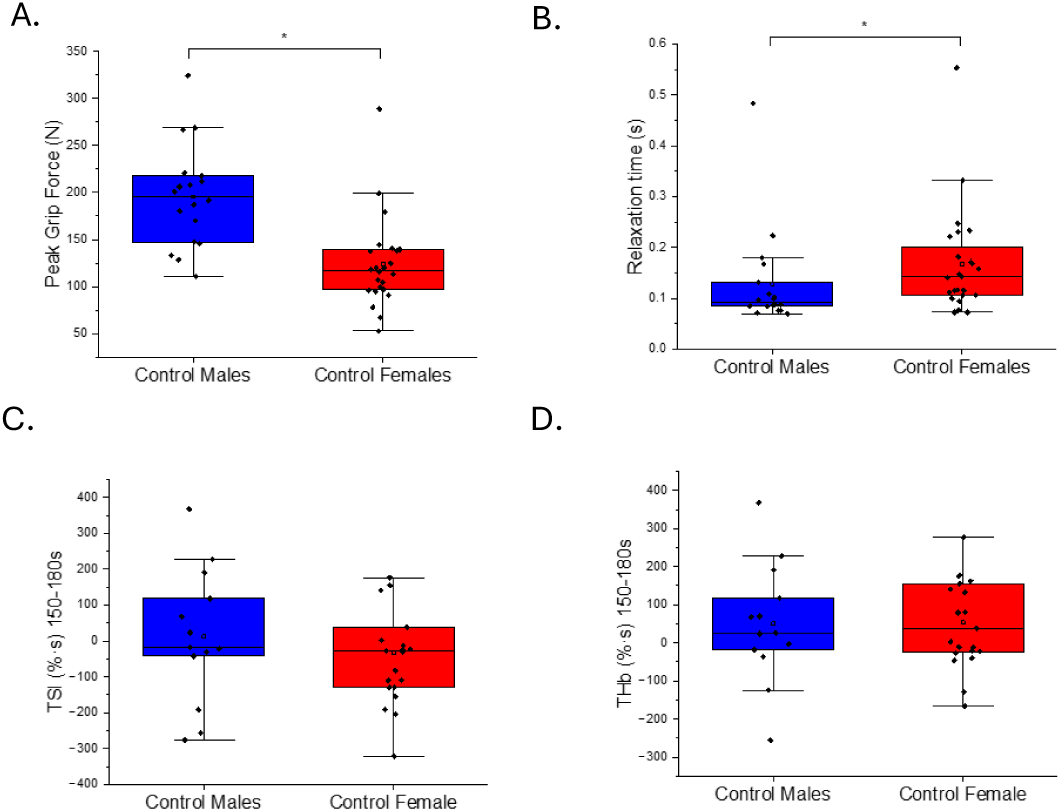
Sex Difference in rHCR and NIRs parameters for combined control groups. a. rHCR derived Peak Grip Force (N). b. rHCR derived Relaxation Time c. Integrated TSI (%·s) for the last 30 seconds of the 180 second hand grip exercise test. d. Integrated THb (µL·s) for the last 30 seconds of the 180 second hand grip exercise test. Data are for MASS Lifecourse and Mitochondrial Dysfunction controls combined. Plots show the mean (empty square). median (centre line), interquartile range (IQR, box), and the most extreme values within 1.5 × IQR (whiskers). Individual data points are shown (blue = male, red =female).

**Figure 5.**
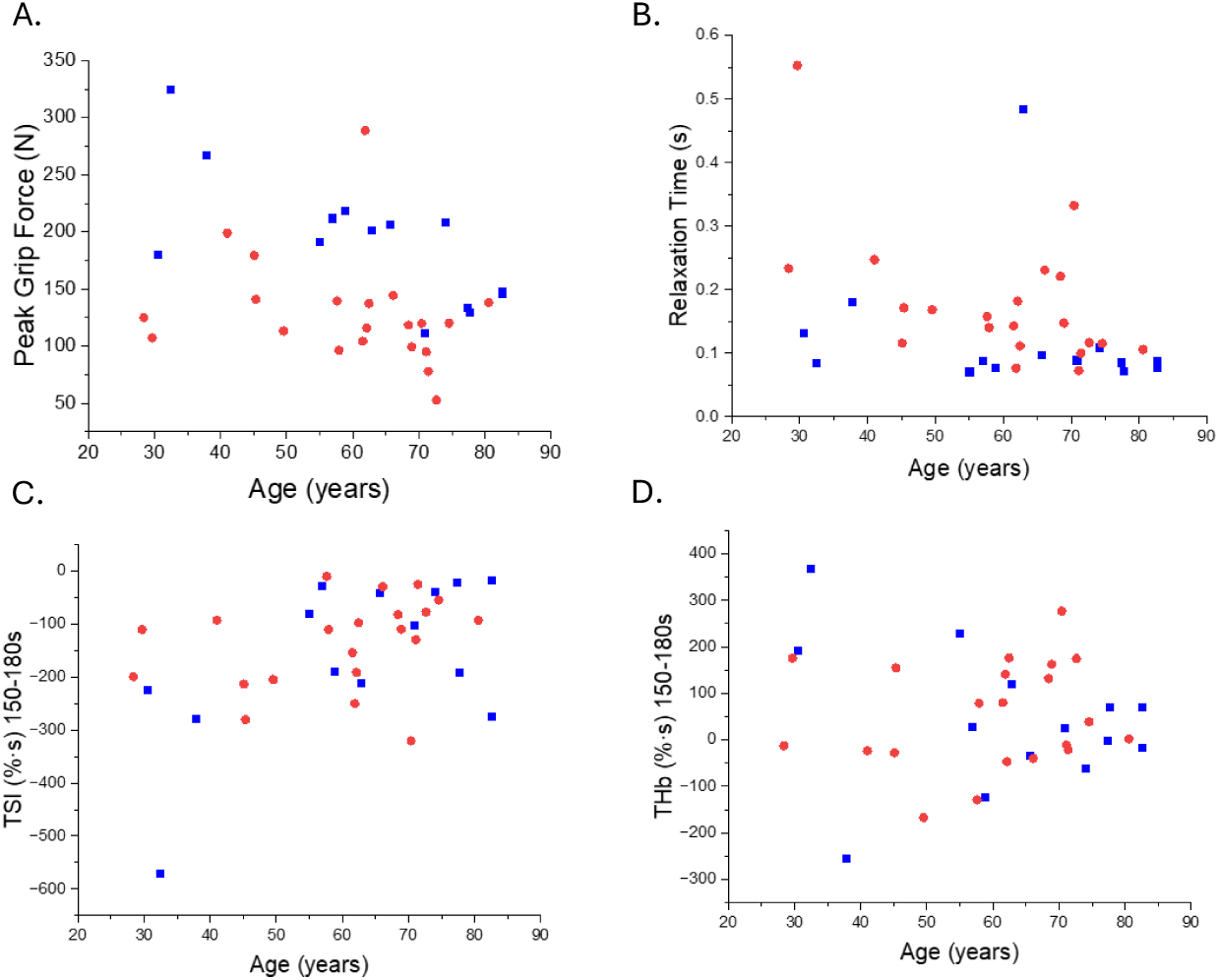
Age Difference a. rHCR derived Peak Grip Force in Newtons (N) plotted according to age. b. rHCR derived Relaxation Time in seconds (s) plotted according to age c. Integrated TSI (%·s) for the last 30 seconds of the 180 second hand grip exercise test plotted according to age. d. Integrated THb (µL·s) for the last 30 seconds of the 180 second hand grip exercise test plotted according to age. Data are for MASS Lifecourse and Mitochondrial Dysfunction controls combined. Males = blue; Females = red.

## Discussion

In this study a brief, non-ischaemic intermittent isometric handgrip exercise protocol combined with NIRS identified divergent muscle oxygenation responses in genetically confirmed mitochondrial disease and sarcopenia. Participants with mitochondrial disease showed an *attenuated* reduction in TSI during exercise, whereas participants with probable or confirmed sarcopenia showed exaggerated reduction in TSI. These opposing responses were observed using the same experimental paradigm. Genetically confirmed mitochondrial disease therefore provided a biologically defined reference phenotype against which the physiological response observed for the more heterogeneous condition of sarcopenia could be interpreted.

### Genetic mitochondrial disease as a reference model of impaired oxidative phosphorylation

As mitochondrial disease provides a biologically validated model of impaired oxidative phosphorylation, it offers a useful reference against which NIRS responses can be interpreted. We show that NIRS performed during a brief, non-ischaemic intermittent isometric and submaximal handgrip exercise detects impaired muscle oxygenation in participants with genetically confirmed mitochondrial disease (**Figure 2c,e**). The absence of a difference in THb suggests that the altered TSI response in participants with genetically confirmed mitochondrial disease was not accompanied by a difference in local blood volume. This extends previous work using a similar forearm exercise paradigm combined with repeated venous blood sampling^12^ and NIRS handgrip protocols involving prolonged arterial occlusion for signal calibration^10,11^. By avoiding arterial occlusion, or repeated venous sampling, the present protocol is potentially more suitable for routine clinical and research use. To explore whether TSI variation reflected disease severity, we examined associations with NMDAS score, exercise intolerance, habitual physical activity and heteroplasmy level, but found no clear relationships, although interpretation was limited by the small, genetically heterogeneous cohort.

### Divergent muscle oxygenation in sarcopenia

In contrast to individuals with genetically confirmed mitochondrial dysfunction, participants recruited from the community with probable or confirmed sarcopenia exhibited an exaggerated reduction in TSI during handgrip exercise (**Figure 6**). This divergent response suggests that altered muscle oxygenation in sarcopenia cannot be explained by the same physiological consequences of impaired oxidative phosphorylation observed in mitochondrial disease. Physical activity, or inactivity, is a recognised confounder when interpreting age-related changes in mitochondrial function^24^. However, in the present study, despite divergent NIRs responses, *both* the mitochondrial disease and the sarcopenia groups were less physically active than their respective comparison groups. This suggests that reduced physical activity alone does not explain the opposing NIRS findings.

**Figure 6.**
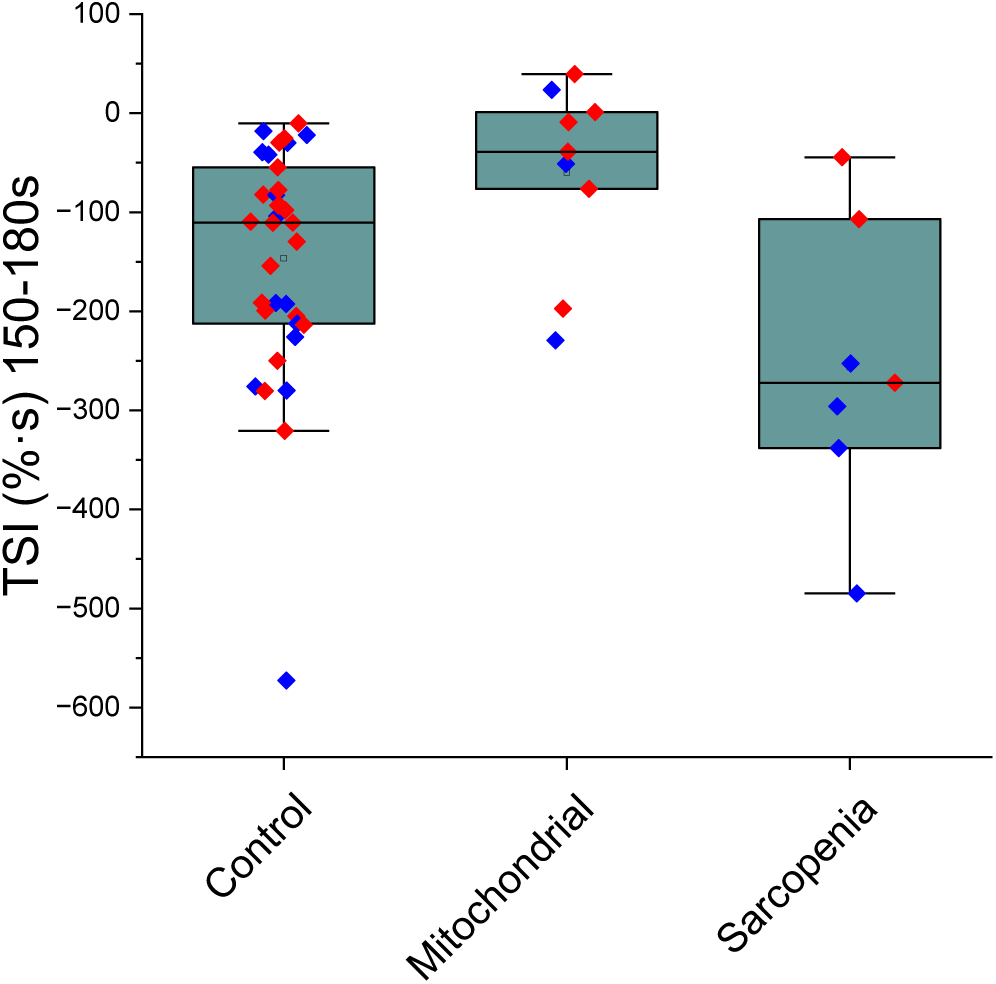
Comparison of Integrated TSI (%·s) for the last 30 seconds of the 180 second hand grip exercise test for participants with genetically confirmed mitochondrial disease (‘Mitochondrial’), probable or confirmed sarcopenia (‘Sarcopenia’) and the combined control cohort. Individual data points are shown (blue = male, red =female). Box-and-whisker plots show the median (centre line), interquartile range (box), and the most extreme values within 1.5 × IQR (whiskers).

One possible explanation for the exaggerated reduction in TSI in sarcopenia is impaired oxygen delivery. Muscle oxygenation reflects the balance between oxygen delivery and utilisation and a greater fall in TSI could also arise from impaired matching of oxygen delivery to metabolic demand. In the current study, THb responses did not differ between groups suggesting that the divergent TSI response was not accompanied by a major difference in local blood volume. However, THb does not directly measure muscle blood flow, and we therefore cannot exclude reduced oxygen delivery as a cause or contributor of exaggerated TSI reduction.

Another possibility is reduced muscle efficiency, resulting in greater oxygen utilisation relative to force production. This interpretation is consistent with MRS studies showing an increased ATP cost of contraction in older and sarcopenic muscle during intermittent isometric exercise, indicating that oxidative phosphorylation remains sufficient to meet demand but requires greater ATP expenditure to generate a given level of force^25–31^. Notably, this phenomenon is less consistently observed during sustained contractions and has been attributed to the additional energetic demands of ionic homeostasis during repeated cycles of membrane depolarisation-repolarisation and Ca²⁺ handling^26^. In keeping with the proposal that ionic dyshomeostasis is a hallmark of skeletal muscle ageing^6^, dynamic exercise studies also demonstrate greater muscle acidification with age^27,29,32^ and in sarcopenic muscle^31^. Consequently, the intermittent contraction-relaxation paradigm used here may be particularly sensitive to detect abnormalities in muscle energetics and ion handling that emerge with ageing in general and in sarcopenia in particular.

An important consideration is that probable sarcopenia was defined by impaired chair rise performance, whereas NIRS measurements were acquired from forearm muscles that showed no significant reduction in grip strength (**Figure 3a**). The exaggerated TSI response may therefore represent a compensatory adaptation within a muscle that is still functioning effectively, potentially reflecting increased energetic demands required to maintain force production despite early pathological change. If confirmed in larger studies, this interpretation would suggest that NIRS may be capable of detecting physiological adaptations preceding overt muscle weakness, supporting its further evaluation as a biomarker in clinical trials.

### Sex related variation in muscle oxygenation and relaxation

The sex differences observed in this study warrant further investigation but should be interpreted cautiously given the small subgroup sizes. While no sex difference in TSI %·s 150-180s was evident in controls (**Figure 4c**), the exaggerated TSI reduction in sarcopenia was largely confined to males (**Figure 3c**), raising the possibility that this effect reflects sex-specific aspects of sarcopenic pathology rather than a general physiological or methodological difference.

### Future work

These findings support further evaluation of non-ischaemic intermittent isometric handgrip with NIRS as a physiological assessment of muscle dysfunction across diverse diseases and populations. Priorities include establishing test-retest reliability, replication in larger cohorts and longitudinal studies to determine whether NIRS responses track progression from probable to confirmed sarcopenia or respond to therapeutic interventions. Mechanistic studies incorporating complementary measures of muscle energetics and perfusion could clarify whether the exaggerated TSI reduction observed in sarcopenia reflects impaired oxygen delivery, increased ATP cost of contraction, or both. Because probable sarcopenia in this study was defined by impaired chair-rise performance rather than reduced grip strength, future work should compare oxygenation signatures in clinically weak and strength-preserved muscles to establish whether the observed response reflects compensation or a broader feature of sarcopenic pathology. These studies will help define the biological basis and clinical utility of the divergent NIRS responses across sarcopenia and mitochondrial disease.

## Conclusion

Using genetically confirmed mitochondrial disease as a biologically defined model of impaired oxidative phosphorylation, we show that a brief, non-ischaemic, intermittent and submaximal handgrip exercise protocol combined with NIRS can distinguish between physiologically distinct forms of muscle dysfunction. While mitochondrial disease was associated with an attenuated TSI response, probable and confirmed sarcopenia showed an exaggerated reduction, indicating that sarcopenic muscle dysfunction does not simply recapitulate the physiological phenotype of impaired oxidative phosphorylation. Coupled with a freely available analytical pipeline, this simple, non-invasive approach has considerable potential for physiological phenotyping and mechanistic stratification in clinical and research settings. More broadly, our findings demonstrate how biologically defined rare diseases can provide a powerful framework for interpreting mechanisms in common heterogeneous age-related conditions.

## Data Availability

All data produced in the present study are available upon reasonable request to the authors

## Acknowledgements

The authors thank all participants who generously gave their time to take part in MASS Lifecourse and the Mitochondrial Dysfunction study. We also acknowledge the work of the MASS Lifecourse study team, which in addition to co-authors AAS, MDW, SH, CH, RC and KS, for the assessments described also included: Jonathan G Bunn, Daniela Carpinelli, Antoneta Granic, Emma Grace Lewis, Claire McDonald, Katie Sloan and Holly E Syddall. We thank the MitoCohort team and participants. We thank GP practices in the North East and North Cumbria Clinical Research Network including Northumbria Primary Care for their support of study recruitment and colleagues who supported study delivery. Finally, the authors acknowledge Dr Richard Dodds who was Principal Investigator of MASS Lifecourse until his death in May 2022.

HD, SP, SH, CH, MDW, AAS, RC and KS acknowledge support from the NIHR Biomedical Research Centre: Newcastle (ref: NIHR203309). MDW, AAS and RC also acknowledge support from the Multiple Long-term Conditions Cross-NIHR Collaboration. MDW acknowledges support from the NIHR Newcastle Clinical Research Facility. KS was funded by an NIHR Clinical Lectureship and acknowledges subsequent support from Research Capability Funding from The Newcastle upon Tyne Hospitals NHS Foundation Trust.

The views expressed are those of the authors and not necessarily those of the NIHR or the Department of Health and Social Care.

## Ethical Standards

All authors comply with the Ethical guidelines for authorship and publishing in the Journal of Cachexia, Sarcopenia and Muscle. All persons gave their informed consent prior to their inclusion in the study and all studies were approved by the appropriate ethics committee and have therefore been performed in accordance with the ethical standards laid down in the 1964 Declaration of Helsinki and its later amendments.

## Abbreviations

ATP: adenosine triphosphate
BMI: body mass index
DXA: dual-energy X-ray absorptiometry
EWGSOP2: European Working Group on Sarcopenia in Older People 2019 (revised) guidelines
GLTEQ: Godin Leisure-Time Exercise Questionnaire
rHCR: Revised HandClench Relaxometer
HHb: deoxygenated haemoglobin
MASS: Muscle Ageing and Sarcopenia Study
MVC: maximal voluntary contraction
MRI: magnetic resonance imaging
MVPA: moderate-to-vigorous physical activity
NIRS: near-infrared spectroscopy
NMDAS: Newcastle Mitochondrial Disease Adult Scale
O₂Hb: oxygenated haemoglobin
RAPA: Rapid Assessment of Physical Activity
rHCR: revised HandClench Relaxometer
ROS: reactive oxygen species
THb: Total haemoglobin
THb µM·s 150-180s: Integrated THb (µM·s) calculated for the last 30-second interval of the 3-minute hand-grip exercise test and normalised to the mean THb during the stable pre-exercise period.
TSI: Tissue Saturation Index ([O₂Hb / (O₂Hb + HHb)] × 100))
TSI %·s 150-180s: Integrated TSI (%·s) values calculated for the last 30-second interval of the 3-minute hand-grip exercise test and normalised to a mean TSI during the stable pre-exercise period.
³¹P-MRS: phosphorus magnetic resonance spectroscopy
5xSTS: Five times chair sit to stand test

